# Laboratory evaluation of SARS-CoV-2 antibodies: detectable IgG up to 20 weeks post infection

**DOI:** 10.1101/2020.09.29.20201509

**Authors:** Louise J. Robertson, Julie S. Moore, Kevin Blighe, Kok Yew Ng, Nigel Quinn, Fergal Jennings, Gary Warnock, Peter Sharpe, Mark Clarke, Kathryn Maguire, Sharon Rainey, Ruth Price, William Burns, Amanda Kowalczyk, Agnes Awuah, Sara McNamee, Gayle Wallace, David Hunter, Steve Segar, Connie Chao Shern, M. Andrew Nesbit, James McLaughlin, Tara Moore

## Abstract

**Background:** The SARS-CoV-2 pandemic necessitated rapid and global responses across all areas of healthcare, including an unprecedented interest in serological immunoassays to detect antibodies to the virus. The dynamics of the immune response to SARS-CoV-2 is still not well understood and requires further investigation into the longevity of humoral immune response that is evoked due to SARS-CoV-2 infection.

**Methods:** We measured SARS-CoV-2 antibody levels in plasma samples from 880 people in Northern Ireland using Roche Elecsys Anti-SARS-CoV-2 IgG/IgA/IgM, Abbott SARS-CoV-2 IgG and EuroImmun IgG SARS-CoV-2 ELISA immunoassays to analyse immune dynamics over time. We undertook a laboratory evaluation for the UK-RTC AbC-19 rapid lateral flow immunoassay (LFIA), for the target condition of SARS-CoV-2 Spike protein IgG antibodies using a reference standard system to establish a characterised panel of 330 positive and 488 negative SARS-CoV-2 IgG samples.

**Results:** We detected persistence of SARS-CoV-2 IgG up to 140 days (20 weeks) post infection, across all three laboratory-controlled immunoassays. On the known positive cohort, the UK-RTC AbC-19 lateral flow immunoassay showed a sensitivity of 97.58% (95.28%-98.95%) and on known negatives, showed specificity of 99.59% (98.53 %-99.95%).

**Conclusions:** Through comprehensive analysis of a cohort of pre-pandemic and pandemic individuals, we show detectable levels of IgG antibodies, lasting up to 140 days, providing insight to antibody levels at later time points post infection. We show good laboratory validation performance metrics for the AbC-19 rapid test for SARS-CoV-2 Spike protein IgG antibody detection in a laboratory based setting.

## Introduction

The World Health Organization declared a pandemic in March 2020 due to severe acute respiratory syndrome coronavirus-2 (SARS-CoV-2), identified late 2019 in Wuhan, China, causing COVID-19 disease (1,2).

A global race ensued to develop diagnostic assays, with the most common being viral RNA detection (RT-qPCR assays), to detect acute infection(3). RT-qPCR assays are labour and reagent intensive, limited by a short temporal window for positive diagnosis, and exhibit potential for false negative results (4). Evidence suggests sensitivity of RT-qPCR can be as low as 70% (5). Lockdown measures and “flattening the curve” strategies meant many infected individuals were instructed to self-isolate and were not offered a diagnostic RT-qPCR, with much of the testing limited to patients admitted to hospital, who perhaps reflect a more severely infected cohort. Consequently, a potentially large number of cases were unconfirmed or undetected(6).

The ability to accurately detect SARS-CoV-2 specific antibodies, which develop after an immune response is evoked, is vital for building biobanks of convalescent sera for treatment, monitoring immune response to infection alongside surveillance studies and assessing responses to vaccination programmes. The timing for when antibody against the novel SARS-CoV-2 virus can be measured is at this time not fully characterised.

Commercial serology immunoassays are mostly laboratory-based and measure IgG antibody levels in plasma or serum. Alternatively, lateral flow immunoassays (LFIAs), require a finger prick blood sample and can be used at point-of-care (POC) or in the home; particularly important in the context of lockdown enforcement during the pandemic. Currently, a limited number of laboratory-based chemiluminescence immunoassays are approved for use in the UK including the Roche Elecsys Anti-SARS-CoV-2 IgG/IgA/IgM against the SARS-CoV-2 Nucleocapsid antigenic region (Roche Diagnostics, Basel, Switzerland) and the Abbott SARS-CoV-2 IgG assay against the same antigenic region (Abbott Diagnostics, Abbott Park, IL, USA).

The complexities of the humoral immune response to SARS-CoV-2 is a much-debated topic. In a US study, approximately one in 16 individuals lacked detectable IgG antibodies up to 90 days post symptom onset, despite previous RT-PCR confirmed infection (7). Patients who remain asymptomatic may mount a humoral immune response which is short-lived, with detectable levels of antibody falling rapidly (8). This, alongside potentially low sensitivity and lack of RT-PCR test availability across the UK has hindered development of well characterised gold standard serology test for IgG antibodies to SARS-CoV-2.

Herein, we describe the use of Roche and Abbott commercial immunoassays, as well as the EuroImmun Anti-SARS-CoV-2 ELISA-IgG against the S1 domain of the spike antigenic protein of SARS-CoV-2 (EuroImmun UK, London, UK) to characterise pre-pandemic and pandemic COVID-19 blood samples (n=880) from within Northern Ireland and report on longevity of IgG antibodies detected. Presently, there is no gold standard assay for comparison, therefore we aimed to establish a reference based on a positive COVID-19 antibody status. We present results of a laboratory evaluation of the UK-RTC AbC-19 with a target condition of antibodies against a cohort of 330 known IgG antibody positive samples according to this ‘positive by two’ system and 488 negative samples (223 pre-pandemic assumed negative and 265 known negative) for IgG to SARS-CoV-2.

## Methods

### Participant samples

The flow of participant samples is summarised in Figure S1. All participants provided informed consent with no adverse events. An online recruitment strategy was employed, with the study advertised through internal Ulster University email, website and social media. A BBC Newsline feature providing the pandemic study email address also prompted interest from the general population.

A small cohort (n=19) of anonymised plasma samples were obtained from a partner USA laboratory for initial protocol development only. The first 800 respondents who expressed interest were provided with an online patient information sheet, consent form and health questionnaire and invited to register to attend a clinic. Exclusion criteria related to blood disorder or contraindication to giving a blood sample. To enrich the cohort for samples potentially positive for SARS-CoV-2 IgG antibody, further participants were invited if they had previously tested PCR positive or had the distinctive symptom of loss of taste and smell. Blood sampling clinics were held at locations around Northern Ireland between April and July 2020 resulting in collection of 263 10ml EDTA plasma samples from 263 separate study participants. Additional anonymised plasma samples were obtained from Southern Health and Social Care Trust (SHSCT) Healthcare workers (n=195), and Northern Ireland Blood Transfusion Service (NIBTS, n=184) through convalescent plasma programs.

Pre-pandemic samples (prior to June 2019, n=136) were obtained from Ulster University ethics committee approved studies with ongoing consent and from NIBTS (n= 200, more than 3 years old). Plasma samples were used at no more than 3 freeze-thaw cycles for all analyses reported within this manuscript.

### Clinical information

Basic demographic information and data with regard to probable or definite prior infection with SARS-CoV-2 virus was obtained from PANDEMIC study participants through the secure online questionnaire requiring responses about positive RT-PCR result and/or time from symptom onset. Anonymised participant samples from USA, SHSCT and NIBTS were provided with age, gender and time since PCR-positive, where a previous test had been carried out.

### Laboratory-based immunoassays

Details of laboratory immunoassays are summarised in supplementary methods and Table S1.

### UK-RTC AbC-19 LFIA

UK-RTC AbC-19 testing was conducted at Ulster University according to manufacturer’s instructions (details in Table S1). Assays were performed as cohorts, with samples in batches of 10, with one researcher adding 2.5µL of plasma to the assay and a second adding 100µL of buffer immediately following sample addition. After 20 minutes, the strength of each resulting test line was scored from 0-10 according to a visual score card (scored by 3 researchers; Figure S2). A score ≥1 was positive. Details of samples used for analysis for detection of antibodies are available in Supplementary methods.

### Statistical analysis

As per Daniel (9) a minimum sample size based on prevalence can be calculated using the following formula: 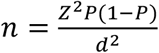, where n = sample size, Z = Z statistic for a chosen level of confidence, P = estimated prevalence, and d = precision. Assuming a prevalence of SARS-CoV-2 of 10% and a precision of 5%, we estimate that the required sample size at 99% confidence (Z = 2.58) to be 240 individuals. If the true prevalence is lower, 5%, the estimated required sample size given a precision of 2.5% is 506 individuals. A minimum sample size of 200 known positives and 200 known negatives is given within MHRA guidelines for SARS-CoV-2 LFIA antibody immunoassays(10).

Statistical analysis was conducted in in R v 4.0.2(11). To assess discordance between test results, data was first filtered to include individuals with an Abbott test result in the range ≥0.25 & ≤1.4, with a 2 × 2 contingency table produced that comprised all possible combinations of [concordant|discordant] test results [within|outside of] this range. A p-value was derived via a Pearson χ^2^ test after 2000 p-value simulations via the stats package.

AbC-19 LFIA performance analyses were performed using MedCalc online (MedCalc Software, Ostend, Belgium). ROC analysis was performed via the pROC package. To compare test result (Positive|Negative) to age, a binary logistic regression model was produced with test result as outcome – a p-value was then derived via χ^2^ ANOVA. To compare time against test result (encoded continuously), a linear regression was performed. We calculated median per time-period and then converted these to log [base 2] ratios against the positivity cut-off for each assay. All plots were generated via ggplot2 or custom functions using base R(12).

## Results

We analysed samples from a mixed cohort of individuals from the general public (n=279), Northern Ireland healthcare workers (n=195), pre-pandemic blood donations and research studies (n=223) and through a convalescent plasma program (n=183). Antibody levels in plasma from these 880 individuals were assessed using the three SARS-CoV-2 immunoassays; EuroImmun IgG, Roche Elecsys IgG/IgM/IgA and Abbott Architect IgG (Table S1). This included a cohort of 223 pre-pandemic plasma samples collected and stored during 2017 to end of May 2019 to determine assay specificity. Of the 657 participants whose samples were collected during the pandemic, 265 (40.33%) previously tested RT-PCR positive with a range of 7-173 days since diagnosis. A total of 225 participants gave time since self-reported COVID-19 symptoms, with a range of 5-233 days from symptom onset, whilst 198 had no symptom or PCR data available.

### Laboratory based antibody immunoassays

A positive result for antibody on one or more of the three laboratory immunoassays was recorded for 385/657 (58.6%) participants who provided a sample during the pandemic. By EuroImmun ELISA, 346 were positive, 20 borderline and 291 were negative. The Roche assay detected 380 positive and 277 negative, whilst Abbott determined 310 positive and 347 negative (Table S2). The median age across all age groups combined was lower for participants testing positive across each of the immunoassays (median [sd] for positive versus negative, respectively: EuroImmun, 41 [13.16] vs 48 [12.95]; Roche, 42 [13.08] vs 48 [13.00]; Abbott, 41 [13.18] vs 47 [13.09]). (Figure S3, p<0.0001). When segregated by age group, however, differences were less apparent in certain groups (Figure S4). Excluding the pre-pandemic cohort, this gap reduced but remained statistically significant EuroImmun, 41 [13.18] vs 45 [12.49]; Roche, 42 [13.15] vs 45 [12.49]; Abbott, 41 [13.26] vs 44 [12.63]) (p<0.01) (median [sd] for positive versus negative). Of note, out of 265 individuals with a previous positive RT-PCR result for SARS-CoV-2 viral RNA, 14 (5.2%) did not show detectable antibodies by any of the three immunoassays, with no association found with age, gender or time between test and blood draw (data not shown).

The three commercial laboratory immunoassays provide a ratio value that increases with IgG antibody titre. When correlation between these values is assessed, good overall agreement is observed between the three immunoassays (Figure 1, Figure S5). As highlighted by Rosadas *et al*., we also see significant disagreement in the Abbott 0.25-1.4 range when compared to EuroImmun and Roche (Figure 1a,b; chi-square p-values: EuroImmun vs Abbott, p<0.001; Roche vs Abbott, p<0.001)(13).

**Figure 1:**
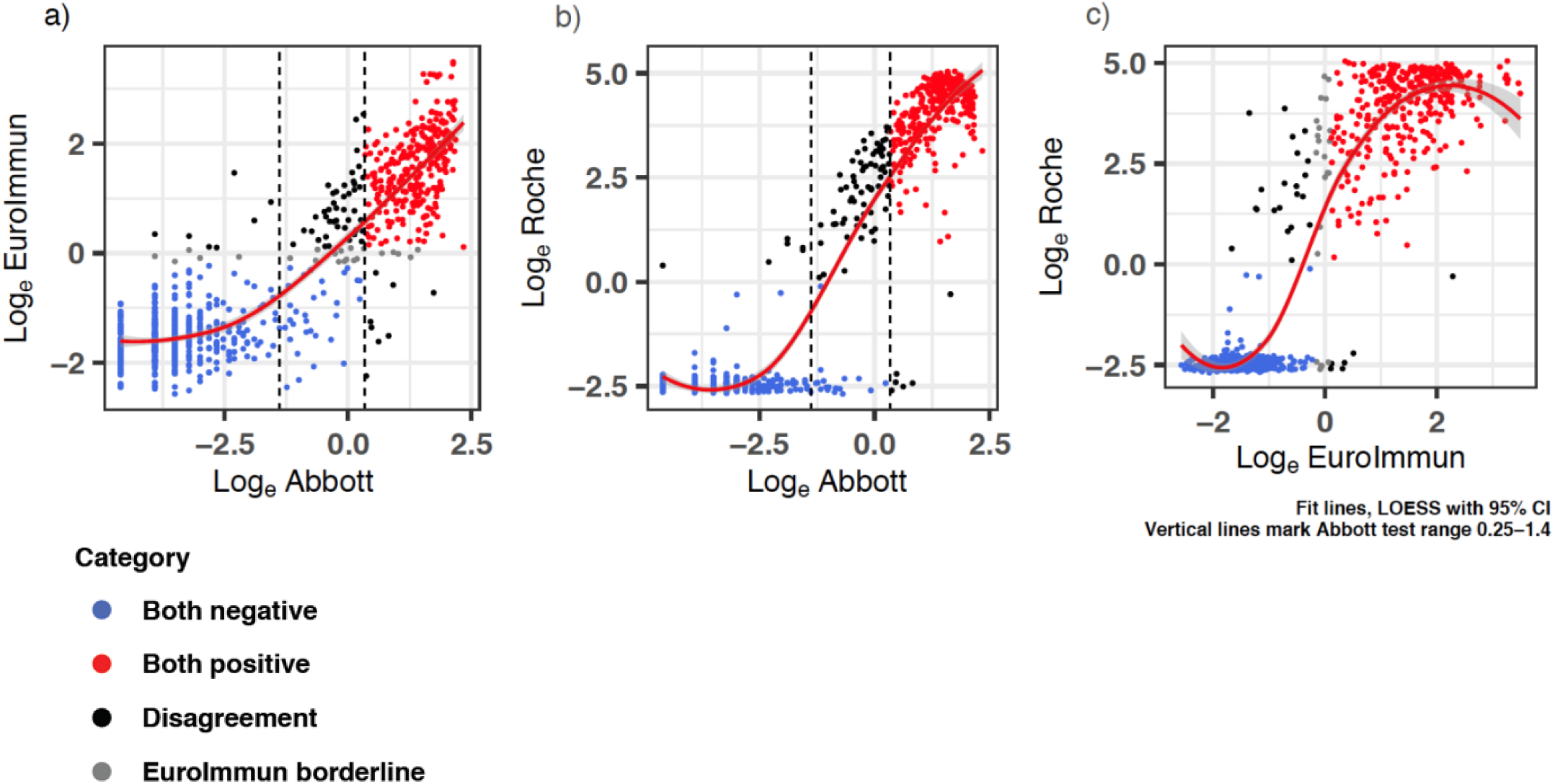
Two-way correlation scatter plots comparing a) EuroImmun b) Abbott and c) Roche immunoassays. Pearson χ^2^ test was used to assess correlations. The results for each test were log transformed to ensure results follow a normal distribution. Negative agreement shown as blue dots, red dots show positive agreement for the two immunoassays, whilst black dots show disagreement and grey dots as the EuroImmun borderline results. Vertical lines mark the Abbott test range 0.25-1.4. n=880. The graphs show positive correlations between all immunoassays evaluated, with the fewest disagreement of results between the Log of Roche and the Log of EuroImmun. Fit lines LOESS, with 95% confidence interval shaded.

### Duration of humoral response to SARS-CoV-2

We found IgG antibodies could still be detected in individuals (excluding pre-pandemic) across all three immunoassays used up to week 20 (day 140) (Figure 2). We note a statistically significant decrease in signal with respect to time across each assay (p-value [slope]): EuroImmun, p=0.036 [-0.785]; Roche, p=0.002 [-0.125]; Abbott, p<0.0001 [-3.585]. These remained statistically significant after adjustment for age. Antibody levels (expressed as a ratio of median result per timepoint divided by positivity cut off; Figure 2d) peaked at Week 1-2 for EuroImmun (1.33) and Abbott (1.64), though reached highest levels at Week 8-12 when measured by Roche (5.45). By week 21-24, median score for all tests had dropped below the positivity cut off, though a small number of RT-PCR positive samples remained above the positive cut off at these later timepoints (Figure 2).

**Figure 2:**
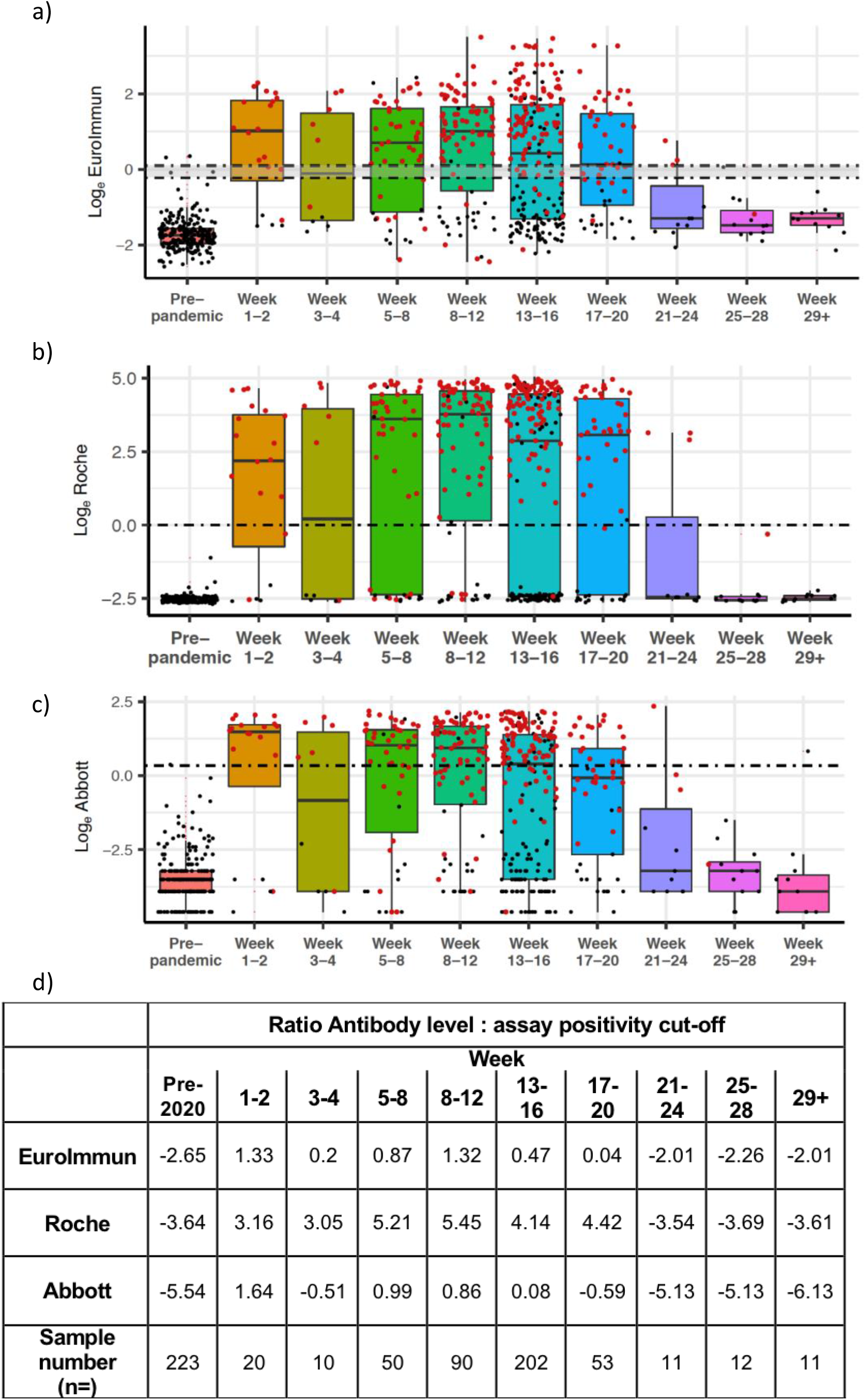
SARS-CoV-2 antibody levels by (a) EuroImmun, (b) Roche, and (c) Abbott, relative to weeks since first reported symptoms or positive PCR result (where data available, n=682). RT-PCR positive individuals are denoted by red dots, while individuals with time since symptom data are denoted in black. Dashed lines delineate loge equivalent of positivity threshold (EuroImmun 1.1, Roche 1.0, Abbott 1.4) for each test, and the negativity threshold for EuroImmun (0.8; borderline result between the two lines). Black bars indicate median, within IQR (interquartile range) boxes for EuroImmun/Roche/Abbott value. Red triangles indicate outliers, based on 1.5* IQR (interquartile range). **(d)** Antibody level ratios for assays over time show varying peaks levels depending on test. Calculated by first establishing the median per time period, then calculating log2 ratio for each period versus each respective assay positivity cut-off.

### UK-RTC AbC-19

Using the commercial immunoassays described we established a well characterised serology sample set of ‘known positive’ and ‘known negative’ for IgG antibodies to SARS-CoV-2 to evaluate performance metrics for the UK-RTC AbC-19 Rapid LFIA.

AbC-19 detects IgG antibodies against the spike protein antigen, so we therefore required all samples to be positive by the EuroImmun SARS-CoV-2 IgG ELISA, which likewise detects antibodies against the S1 domain (14). To develop this characterised cohort, samples were also required to be positive by a second immunoassay (Roche or Abbott). To analyse specificity of the AbC-19 LFIA for detection of SARS-CoV-2 IgG antibody, we assessed 350 plasma samples from participants classed as ‘known negative for SARS-CoV-2 IgG antibody’ on the AbC-19 LFIA. All samples were from individuals confirmed to be negative across all three laboratory assays (Roche, EuroImmun, Abbott). Using these positive n=304 and negative n=350 antibody cohorts, we determined a sensitivity for detecting SARS-CoV-2 IgG antibody of 97.70% (95% CI; 95.31%-99.07%) and specificity of 100% (98.95%-100.00%) for the AbC-19 LFIA (Table 1).

**Table 1:** UK-RTC AbC-19 LFIA performance metrics against known antibody positive and known antibody negative cohorts.

| Total Negative | True Negative | False Positive | Total Positive | True Positive | False Negative | Sensitivity % (95 CI) | Specificity % (95 CI) |
| --- | --- | --- | --- | --- | --- | --- | --- |
| Pre-pandemic (n=223) |  |  |  |  |  |  |  |
| 223 | 222 | 1 | n/a | n/a | n/a | n/a | 99.55%<br>(97.53% to 99.99%) |
| Initially reported cohorts (n=654) |  |  |  |  |  |  |  |
| 350 | 350 | 0 | 304 | 297 | 7 | 97.70%<br>(95.31%-99.07%) | 100.00%<br>(98.95%-100.00%) |
| Extended cohorts (n=818) |  |  |  |  |  |  |  |
| 488 | 486 | 2 | 330 | 322 | 8 | 97.58%<br>(95.28%-98.95%) | 99.59%<br>(98.53%-99.95%) |

Given a recent report of lower specificity in the AbC-19 LFIA (15) and the possibility of introducing sample bias, we revised our inclusion criteria for the negative cohort. For the pre-pandemic cohort, we included samples from all 223 individuals, regardless of results on other laboratory immunoassays. When this assumed negative pre-pandemic cohort was used for laboratory evaluation for target condition of antibodies, we observed a specificity of 99.55% (97.53% to 99.99%, Table 1). We obtained more AbC-19 devices and expanded the negative cohort to include all samples that matched our criteria (samples collected during the pandemic to be negative by all three laboratory assays and all pre-pandemic samples regardless of other immunoassay results). The specificity observed on this extended negative cohort of 488 samples was 99.59% (98.53% to 99.95%, Table 1). For sensitivity analysis on a positive cohort (samples positive by EuroImmun and one other test), we were able to analyse all samples previously untested due to limited testing capacity and tested a positive cohort of 330 samples giving a sensitivity of 97.58% (95.28% to 98.95%, Table 1).

When used for its intended use case, the AbC-19 LFIA provides binary positive/negative results. However, when assessing LFIA in the laboratory, each test line was scored against a scorecard by three independent researchers (0 negative, 1-10 positive; Figure S2). Compared to quantitative outputs from the Abbott, EuroImmun and Roche assays, the AbC-19 LFIA shows good correlation (Abbott r=0.84 [p<0.001]; EuroImmun r=0.86 [p<0.001]; Roche r=0.82 [p<0.001]; Figure 3, Figure S5-Figure S7).

**Figure 3:**
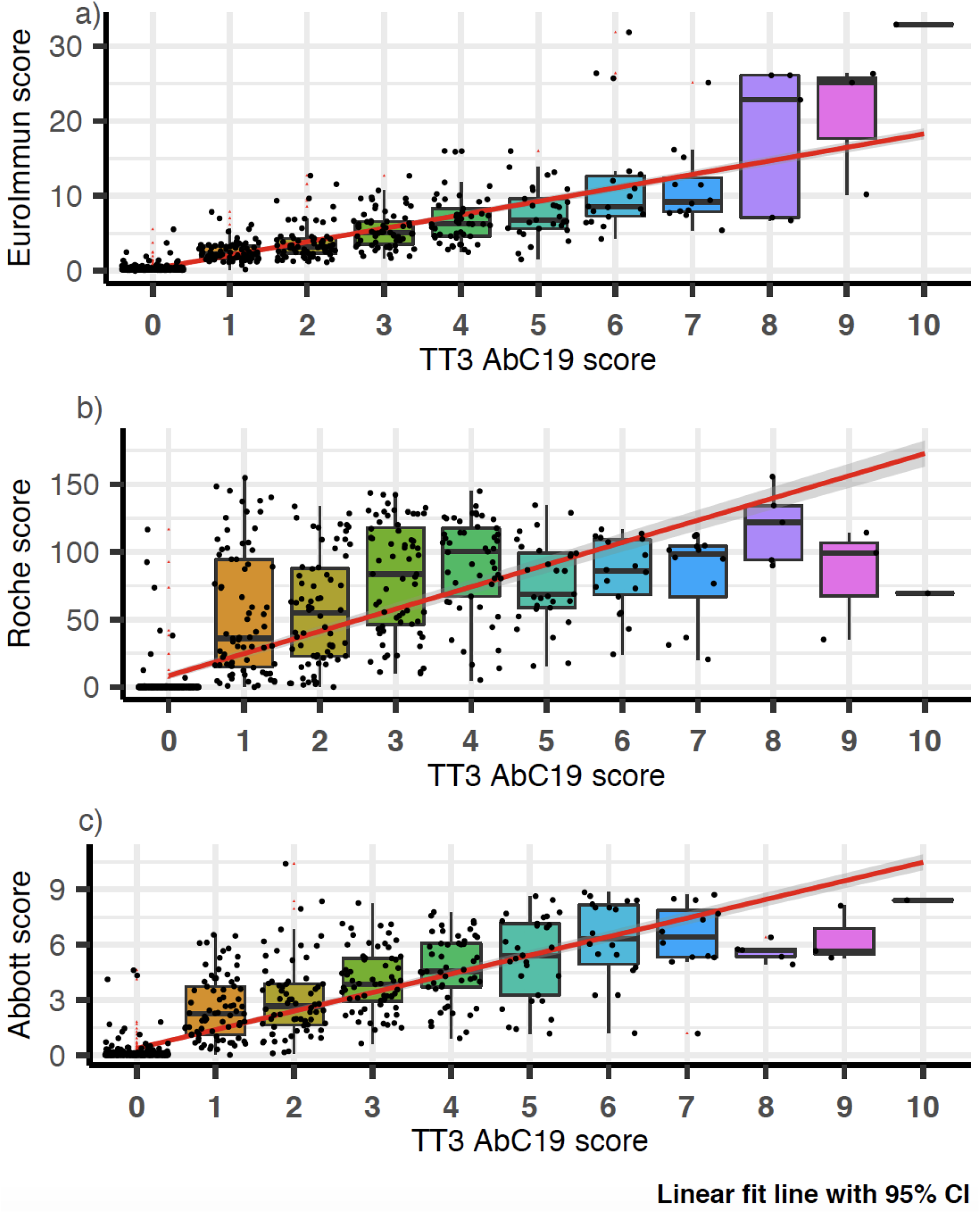
AbC-19 extended cohort (n=818) correlation to a) EuroImmun b) Roche and c) Abbott scores. Box plots overlaid on scatter plot, comparing AbC-19 test scores to EuroImmun, Roche and Abbott quantitative antibody values. Red linear line of best fit with 95% confidence interval shaded in grey. Black bars indicate median, within IQR (interquartile range) boxes for EuroImmun/Roche/Abbott value. Red triangles indicate outliers, based on 1.5* IQR (interquartile range).

### Analytical specificity and sensitivity of AbC-19 LFIA

We observed no cross-reactivity across samples with known H5N1 influenza, Respiratory syncytial virus, Influenza A, Influenza B, Bordetella Pertussis, Haemophilus Influenzae, Seasonal coronavirus NL63 and 229E on the AbC-19 LFIA (n=34 samples, n=8 distinct respiratory viruses; Table S3). Against a panel of external reference SARS-CoV-2 serology samples, the AbC-19 LFIA detected antibodies with scores commensurate to the EuroImmun ELISA scores (Figure S8, Table S4).

## Discussion

Serological antibody immunoassays are an important tool in helping combat the SARS-CoV-2 pandemic. One difficulty faced in validation of antibody diagnostic assays has been access to samples with known SARS-CoV-2 antibody status. As previously described, there is no clear gold standard for reference against which to assess SARS-CoV-2 immunoassays. A positive RT-PCR test has been used previously to indicate previous COVID-19 infection, though this approach is limited by a high rate of false negatives, failure in some cases to develop IgG antibodies (sero-silence or lack of antibody against the same antigenic component of the virus as the immunoassay uses as a capture antigen) and the lack of RT-PCR testing availability early in the pandemic (3,5,16). We failed to detect SARS-CoV-2 IgG antibody in 14 of 265 (5.2%) of previously RT-PCR SARS-CoV-2 viral RNA positive participants in this study. It is unclear if this is due to insufficient/absent antibody production in these individuals, or due to a false positive PCR result which may occur in the UK at a rate between 0.8-4.0% (17). Self-assessment of symptoms for COVID-19 disease is a poor indicator of previous infection, even amongst healthcare workers (18). Asymptomatic individuals may be unaware of infection and others may harbour pre-existing immunity or elucidate a T cell response. Additionally, the kinetics of a SARS-CoV-2 virus infection contributes to the loss of sensitivity of RT-PCR to detect virus with time, contributing to false negative RT-PCR test results for individuals who may be late to present for virus detection tests (5,19).

Our results show strong correlation between all three immunoassays, with shortcomings in the Abbott system output 0.25-1.4 range, as described previously, suggesting an overestimated positive cut-off (Figure 1) (13). Our detection of antibodies 140 days after RT PCR positive status (20 weeks, and beyond in a small number of samples) indicates persistence IgG antibodies to both the spike protein and nucleocapsid protein, despite typical patterns of antibody decay after acute viral antigenic exposure being as rapid (20). Others have reported SARS-CoV-2 antibodies decline at 90 days (19), we also noted a statistically significant decline over time but levels remain detectable at 140 days (Figure 2). We note that IgG levels reach their peak (Roche ratio 5.45 times threshold cut-off) as late as Week 8-12 from first symptoms or a viral RNA RT-PCR positive result, though this may be an artefact of lower number of participants at earlier timepoints (Figure 2d). Longitudinal studies on SARS-CoV-1 convalescent patients suggests that detectable IgG can still be present as long as 2 years after infection (21). Further studies are needed on large cohorts with sequential antibody immunoassays performed on symptomatic and non-symptomatic individuals as well as those with mild or severe COVID-19 to fully elucidate the humoral immune response to SARS-CoV-2. This is vital to inform vaccine durability, so-called ‘immune passports’ and in the definition of a protective threshold for anti-SARS-CoV-2 antibodies.

To assess sensitivity and specificity of the AbC-19 LFIA for its ability to detect SARS-CoV-2 antibody in a laboratory evaluation, we developed a reference standard for SARS-CoV-2 antibodies, which does not rely on a single test as reference. A similar approach was used in a recent seroprevalence study in Iceland, whereby two positive antibody results were required to determine a participant sample as positive for SARS-CoV-2 antibody (16).

Our evaluation of performance metrics for the UK-RTC AbC-19 LFIA to detect antibodies for SARS-CoV-2 gave 97.58% sensitivity and 99.59% specificity. In a recent evaluation of the AbC-19 tests, Mulchandani et al. observed a specificity of 97.9% (97.2%-98.4%) on a cohort of pre-pandemic samples and report a sensitivity of 92.5% (88.8% to 95.1%) for detecting previous infections (based on a previous RT-PCR result) or 84.7% (80.6% to 88.1%) against the Roche Elecsys antibody test, which detects IgM/IgG/IgA SARS-CoV-2 antibodies to the nucleocapsid portion of SARS-CoV-2 (18).

In our study, good correlation was observed in quantitative score between results on all immunoassays with the highest observed between EuroImmun and AbC-19 LFIA (Figure S6, S7). This is to be expected, given both the AbC-19 LFIA and EuroImmun ELISA detect IgG antibodies against spike protein. For the assessment of immunity to prior natural infection as well as to immunisation, it is important to note IgG antibodies against SARS-CoV-2 spike protein detected by laboratory-based EuroImmun ELISA and AbC-19 LFIA are known to correlate with neutralizing antibodies, which may confer future immunity (22,23).

Previous evaluations of the sensitivity and specificity reported by Public Health England (PHE), showed a EuroImmun sensitivity of 72% and specificity of 99%, Abbott with sensitivity of 92.7% and specificity of 100% and Roche with sensitivity of 83.9% and specificity of 100% (24–26). The PHE analyses for each of these tests used previous infection (RT-PCR positive status) as a reference standard, the limitations of which are discussed above.

In the use of characterised ‘known positive’ and ‘known negative’ cohorts, one limitation of this study is its potential for spectrum bias, whereby our positive-by-two reference system may artificially raise the threshold for positive sample inclusion, possibly resulting in the overestimation of the sensitivity of any test evaluated (27). However, similar issues have been raised when using previous RT-PCR result or definitive COVID-19 symptoms as inclusion criteria given these will likely skew a cohort towards more severe disease (5). Importantly, our mixed origin of samples forming the cohort provides a positive cohort for assessing assay sensitivity that includes individuals from the general public, healthcare workers and from convalescent plasma programmes. Our analysis of specificity on only pre-pandemic individuals (n=223) shows similar specificity (99.55%) to the larger mixed ‘known negative cohort’ (n=488, sensitivity 99.59%). In the absence of a clear gold standard test, our system relies on no single test (each with their individual shortcomings) and instead takes an average of three.

Our assessment of the UK RTC AbC-19 LFIA using our characterised cohorts of known SARS-CoV-2 antibody positive and antibody negative plasma, in a laboratory setting shows good performance metrics for its ability to detect SARS-CoV-2 IgG antibody. We note it uses plasma from venous blood samples, as opposed to the use of a finger prick blood sample. Additionally, when this UK RTC AbC-19 LFIA was used on our cohort, a number of the positive results scored low, (1/10 using the score card under laboratory conditions, Figure 3) with a faint test band visible to a trained laboratory scientist but perhaps difficult to identify as positive by individuals performing a single test (Figure S6). This faint line may be reflective of the longer time from infection for the Northern Ireland cohort used. If this AbC-19 LFIA is to be used in clinical settings it is important to determine if all users observe the same results as observed in this laboratory evaluation.

This assessment of the AbC-19 LFIA does not provide data on how this test will perform in a seroprevalence screening scenario, but instead provides metrics for the performance of the test, where presence of SARS-CoV-2 antibodies is of interest, as opposed to previous COVID-19 infection. An important potential use of the AbC-19 LFIA would be in monitoring the immune response to vaccination, with most vaccines utilising SARS-CoV-2 Spike protein antigens (28). It is not yet known if presence of SARS-CoV-2 antibodies indications immunity from infection.

## Conclusion

We present a comprehensive analysis of 880 pre-pandemic and pandemic individuals and show IgG antibodies are detectable up to 140 days from symptoms or positive RT-PCR test, showing persistence of immunity at later time points than previously published. We use antibody positive as an alternative to RT-PCR positive status as a standard for assessing SARS-CoV-2 antibody assays and show strong performance for the UK-RTC AbC-19 LFIA rapid point of care test in detecting SARS-CoV-2 antibodies. It is fully understood that user experience in future studies in the real world is important and may alter the performance characteristics. Also, the effect of operator training will have direct effects upon test performance. We welcome further clinical evaluation of the AbC-19 LFIA in large cohorts of symptomatic and asymptomatic individuals alongside large studies assessing COVID-19 outcomes in individuals with longitudinal studies to fully validate its implementation across all intended use cases.

## Supporting information

STARD checklist

## Data Availability

The datasets used and/or analysed during the current study are available from the corresponding author on reasonable request.

## Declarations

### Ethics approval and consent to participate

All study participants provided informed consent. This study was approved by Ulster University Institutional Ethics committee (REC/20/0043), South Birmingham REC (The PANDEMIC Study IRAS Project ID: 286041Ref 20/WM/0184) and adhered to the Declaration of Helsinki and Good Clinical Practice.

### Consent for publication

Not applicable.

### Authors’ contributions

TM, JML conceived the study. LR, JM and TM performed all laboratory analyses. LR, SM and KYN analysed data, KB performed all statistical analyses/interpretations and produced figures. NQ, FJ, GW and PS performed all Roche analyses and provided SHSCT cohort samples. MC, KM and SR performed all Abbott analyses and provided Blood Transfusion cohort samples. TM, RP and AN coordinated participant recruitment, consent and sampling. WB and JML developed online consent forms, questionnaires and databases. LR, JM, AK, AA, GW, DH, SS, CCS performed sample collection and processing. LR and TM wrote the manuscript, with significant contributions from JM and KB. All authors reviewed and approved the final manuscript.

## Acknowledgements

We are extremely grateful to all the people of Northern Ireland who took part in this study and gave blood during the pandemic. We are indebted to the phlebotomists-Geraldine Horrigan and Pamela Taylor who conducted the blood draws whilst ensuring the highest possible level of safety to the participants. We are also grateful to Kingsbridge Private Hospital Group for sponsorship and providing everything needed for blood collection including the clinical rooms. We acknowledge Dr Tony Byrne for use of his laboratory and Professor Gareth Davison for laboratory space and equipment during the pandemic within a locked down University.

