## Supplementary Materials for "Laboratory evaluation of SARS-CoV-2 antibodies: detectable IgG up to 20 weeks post infection"

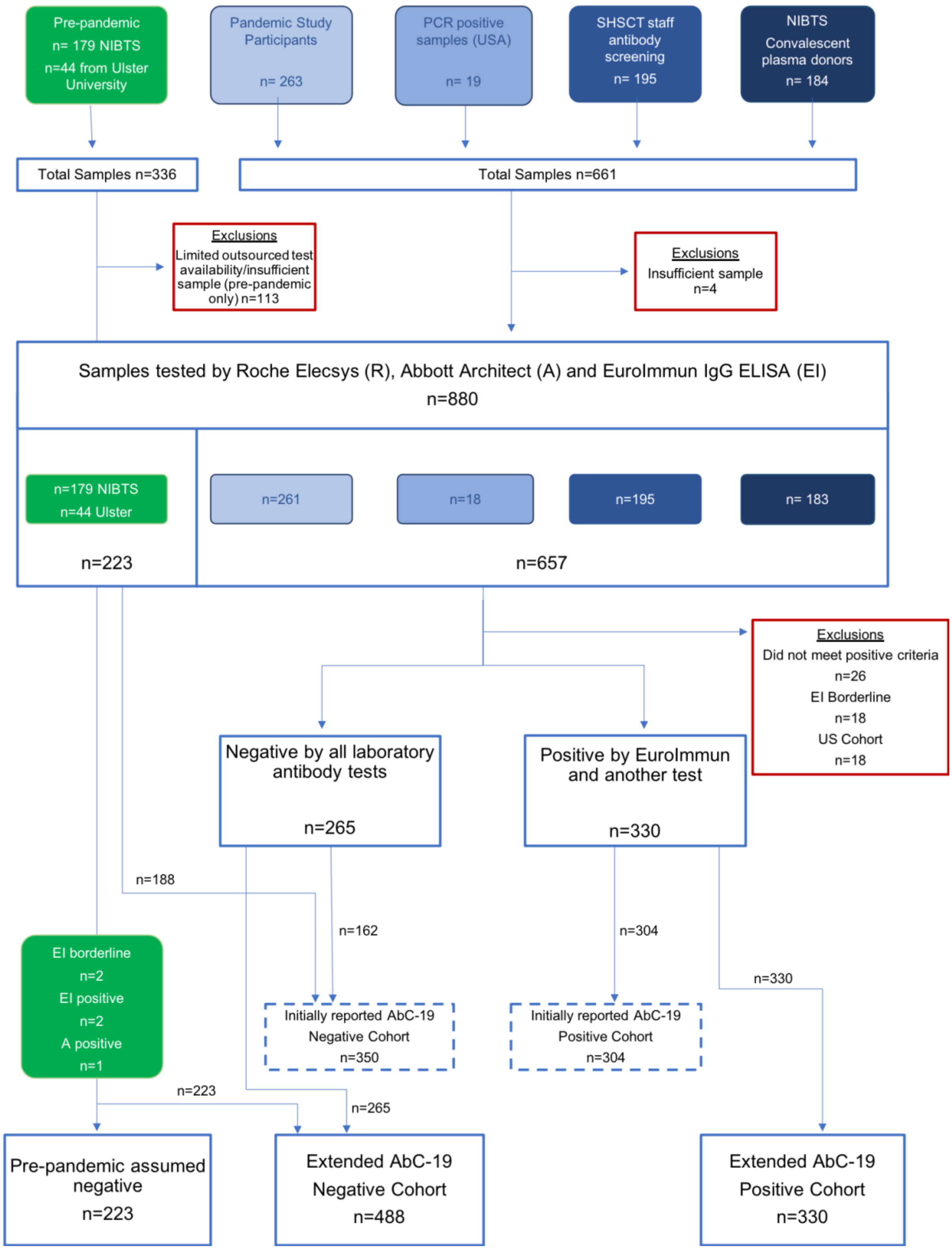

**Figure S1: Flow of participant plasma samples through the study.**

All available samples from participants within each cohort, and the included and excluded samples at all stages. Freeze thaw cycles were closely monitored for all sample aliquots. Pre-pandemic samples taken forward for Roche, Abbott and EuroImmun testing were selected based on aliquot volume and availability.

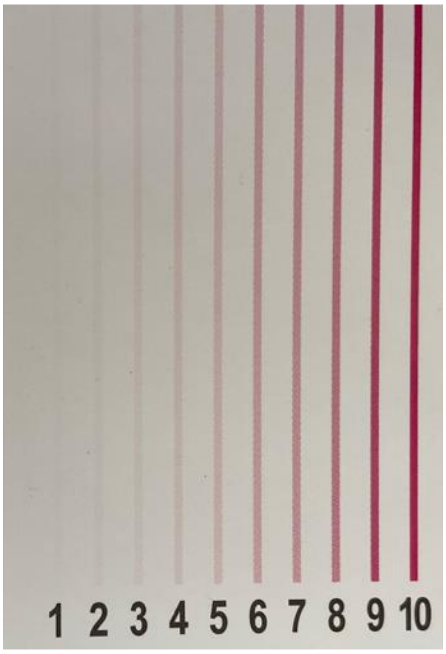


**Figure S2: Visual Score card for quantitative interpretation of AbC-19 LFIA test bands.** A scale of 0 (not pictured, negative-no test line visible) to 10 (positive-strongest test line). Any LFIA scoring 1 or above was classified as positive.

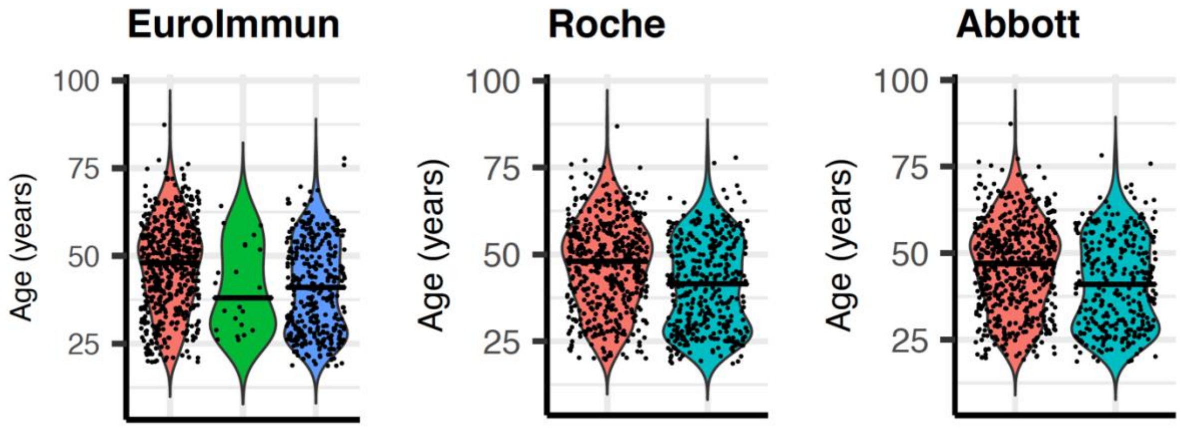


**Figure S3: Age violin plots overlaid with scatter for samples included in correlation analysis (where age data available) n=880.**

The above graphs allow comparison of the distributions and probability density of ages for EuroImmun, Roche and Abbott immunoassays. Wider areas of the violin plot represent high probability density, whilst narrow areas represent low probability density. Horizontal bar indicates median age. The red violin plots represent the negative results, the green violin plot represent the borderline results and the blue/turquoise violin plots represent the positive results.

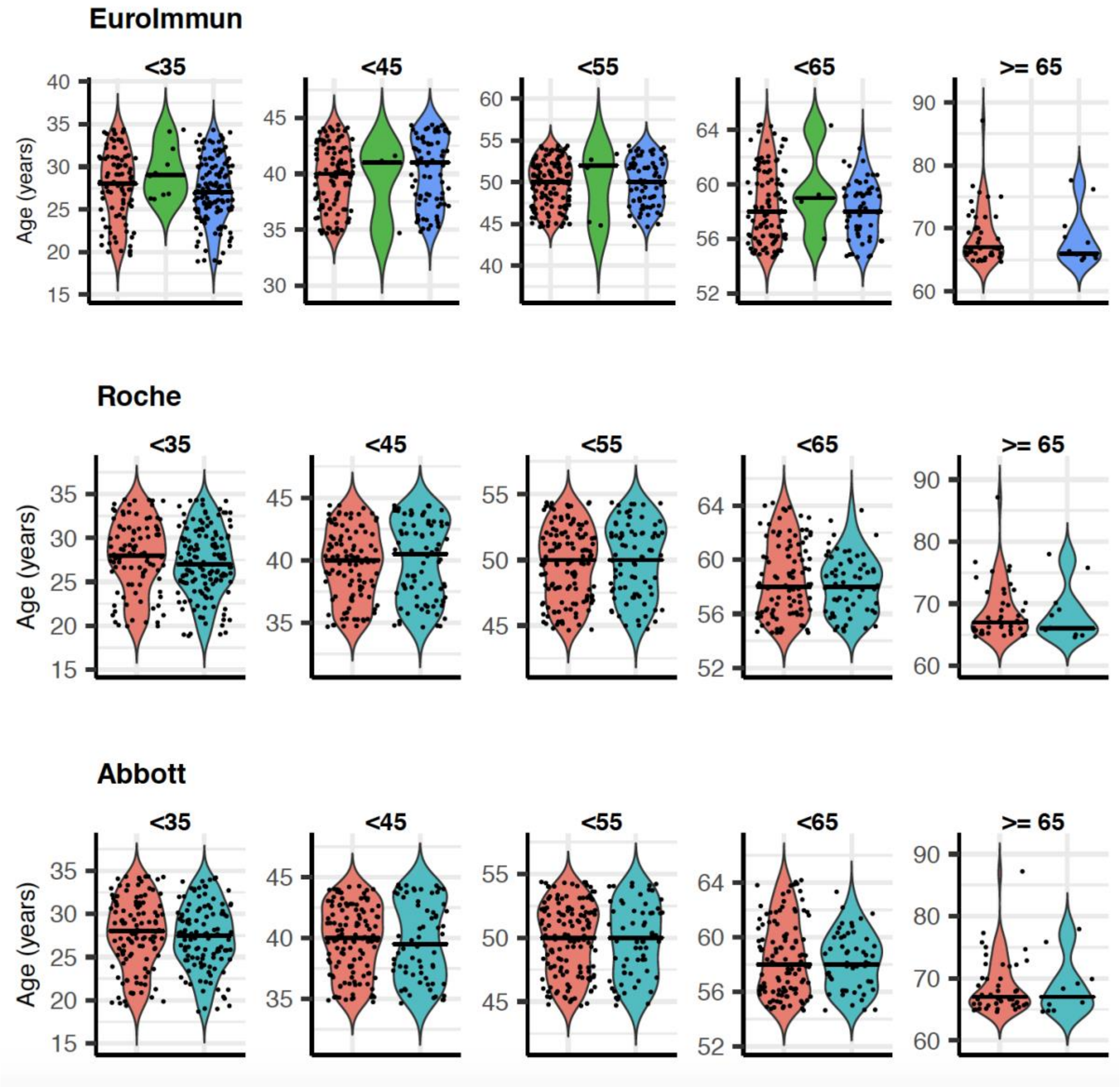


**Figure S4: Age violin plots separated into age groups (where age data available) for samples included in correlation analysis.**

The above figure presents graphs for each immunoassay (EuroImmun, Roche and Abbott) with the corresponding age groups <35 years, <45 years, <55 years, <65 years and >= 65 years. The red violin plots represent the negative results, the green violin plot represents the borderline EuroImmun results, and the blue/turquoise violin plots represent the positive results (n=848).

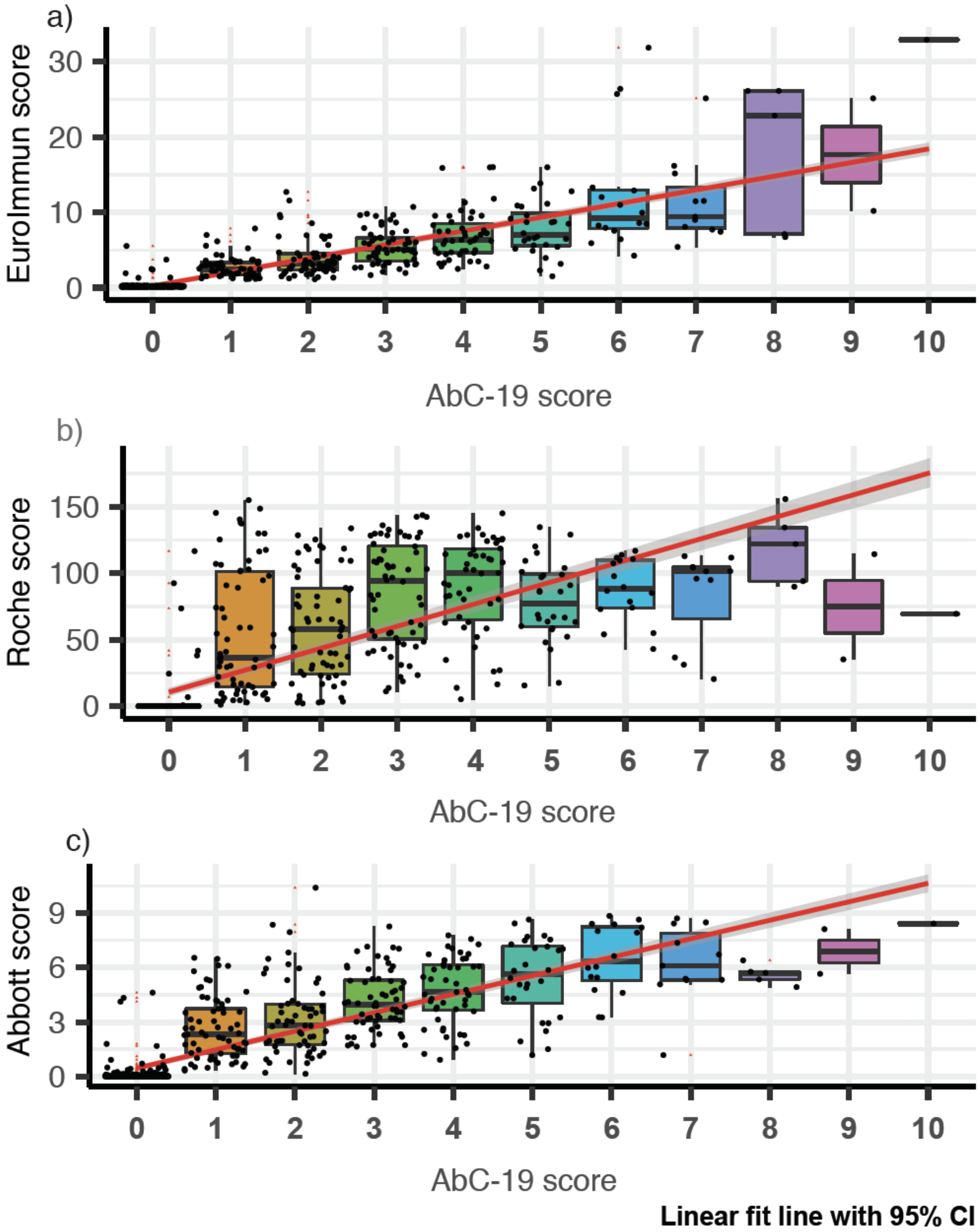


**Figure S5: AbC-19 initially reported cohort n=654 correlation to a) EuroImmun b) Roche and c) Abbott scores.** Box plots overlaid on scatter plot, comparing AbC-19 test scores to EuroImmun, Roche and Abbott quantitative antibody values. Red linear line of best fit with 95% confidence interval shaded in grey. Black bars indicate median, within IQR (interquartile range) boxes for EuroImmun/Roche/Abbott value. Red triangles indicate outliers, based on 1.5* IQR (interquartile range).

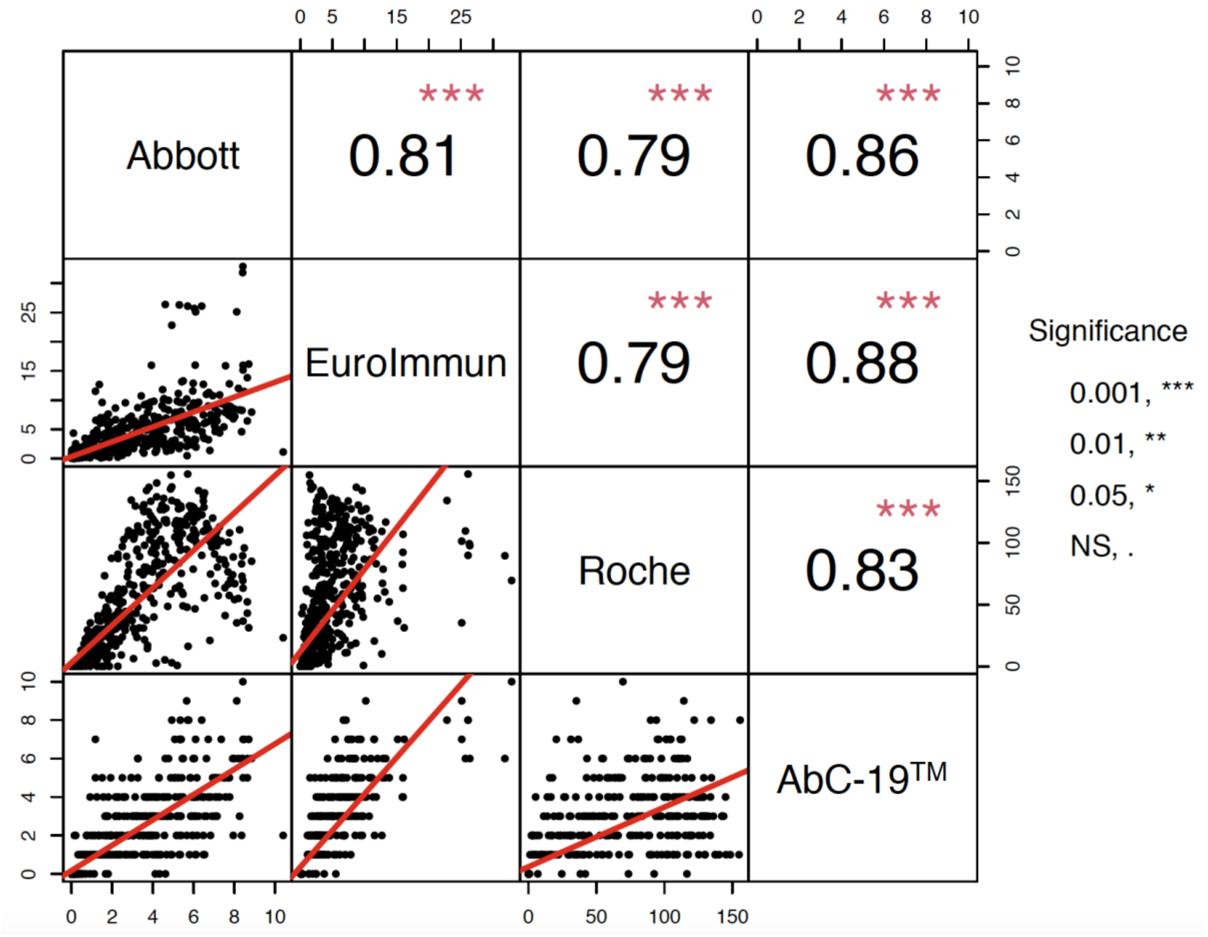


**Figure S6**: **Correlation matrix between Abbott, EuroImmun, Roche and initially reported AbC-19 cohort (n=654) quantitative output values for SARS-CoV-2 antibody levels.** Strong correlations are observed between all immunoassays. The level of significance was set at p<0.05. All immunoassays were significantly correlated p<0.001.

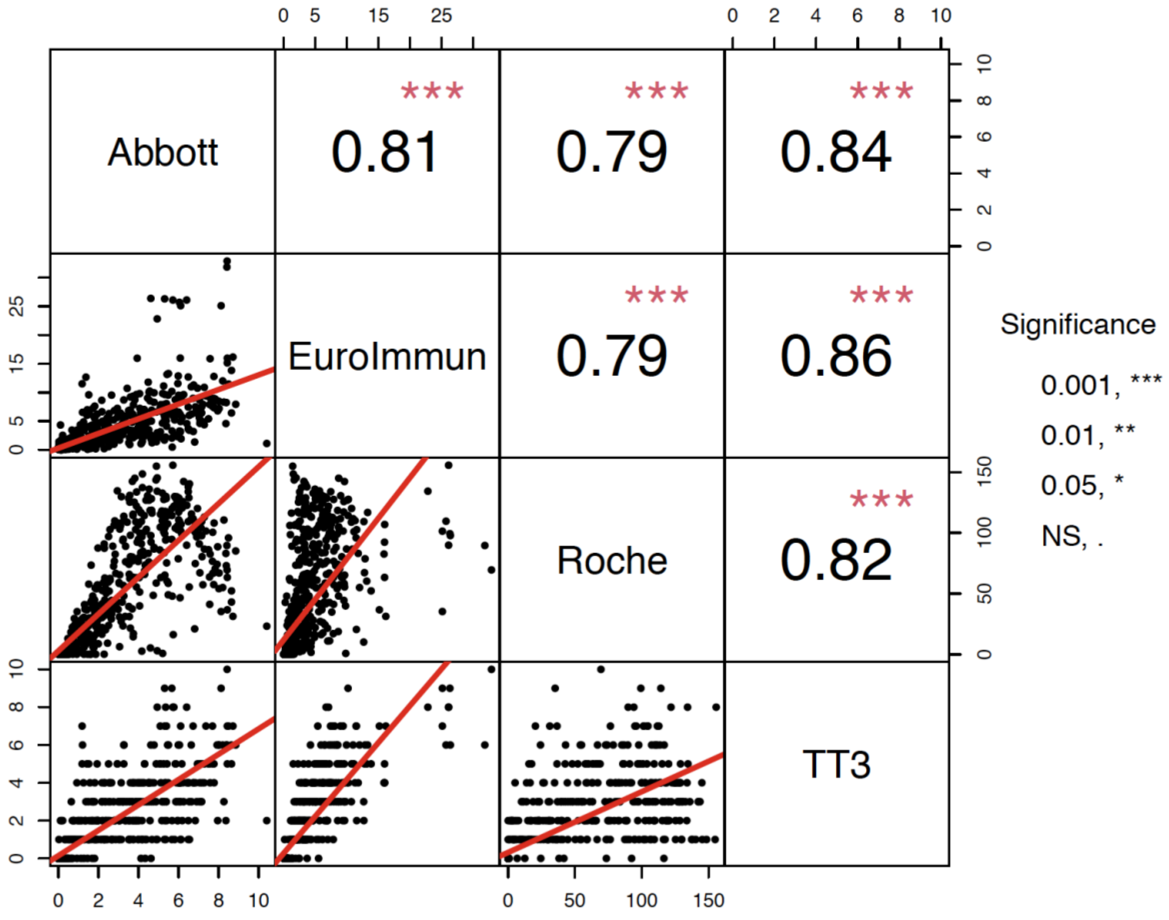


**Figure S7**: **Correlation matrix between Abbott, EuroImmun, Roche and extended AbC-19 cohort (n=818) quantitative output values for SARS-CoV-2 antibody levels.** Strong correlations are observed between all immunoassays. The level of significance was set at $p<0.05$. All immunoassays were significantly correlated $p<0.001$.

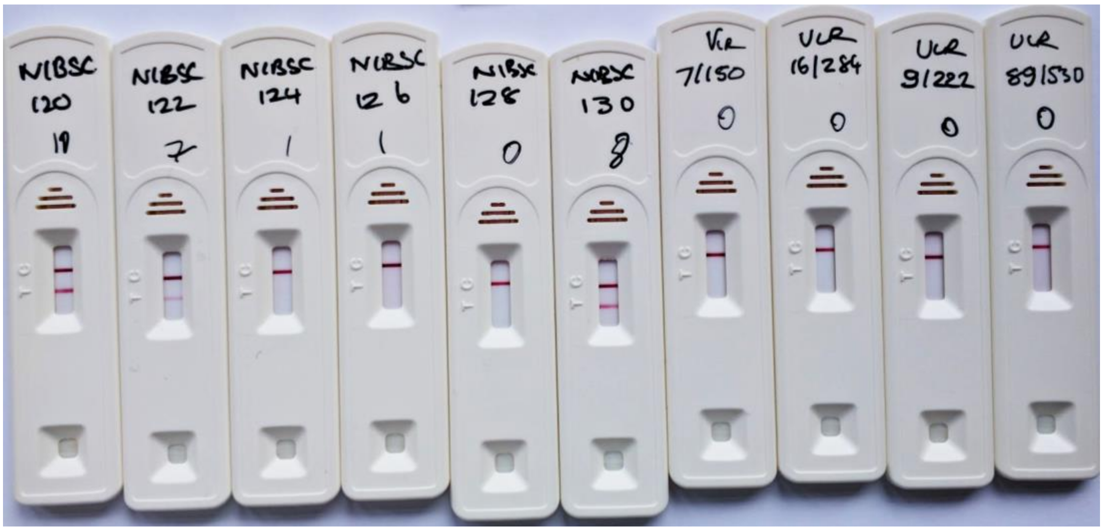

**Figure S8: NIBSC external reference serology standards and known respiratory virus serology samples.**

The scorecard score 0-10 was annotated on test cassette beneath sample ID when agreed by three independent experienced researchers. All LFIAs had a visible control line.

### Table S1: Summary specifications for SARS-CoV-2 immunoassays investigated.

| Immunoassay | Principle | Antigen Target | Assay Time (min) | Antibody Detected | Measurement | Result | Calibration | Evaluation of results | Results |
|---|---|---|---|---|---|---|---|---|---|
| **EuroImmun ELISA** | Enzyme-linked immunosorbent assay (enzyme-HRP) | S1 domain of the spike protein | 120 | IgG | Photometric measurement of the color intensity using wavelength of 450 nm and a reference wavelength between 620 nm and 650 nm | OD (Optical density) | One Positive calibrator | OD of clinical sample/OD of calibrator | < 0.8 Negative, ≥ 0.8 to <1.1 Borderline, ≥ 1.1 Positive |
| **Roche Elecsys immunoassay** | Electro-chemiluminescence | Nucleocapsid | 18 | IgG, IgA and IgM | Application of a voltage to the electrode then induces chemiluminescent emission which is measured by a photomultiplier | RLU (Relative Light Intensity) | One Positive calibrator and one Negative calibrator | The analyzer automatically calculates the cut-off based on the measurement of ACOV2 Cal1 (negative) and ACOV2 Cal2 (positive). The result of a sample is given either as reactive or non-reactive as well as in the form of a cut-off index (COI; signal sample/cut-off). | < 1.0 Negative, ≥ 1.0 Positive |
| **Abbott Architect SARS-CoV-2** | Chemiluminescent microparticle immunoassay | Nucleocapsid | 30 | IgG | The resulting chemiluminescent reaction is measured as a relative light unit (RLU). | RLU (Relative Light Intensity) | One Positive calibrator | Results are reported by dividing the sample result by the calibrator result (mean of 3 calibrators). The default result unit for the SARS-CoV-2 IgG assay is Index (S/C). | < 1.4 Negative, ≥ 1.4 Positive |
| **AbC-19** | Rapid Point of Care Lateral Flow Immunoassay | Full length Spike protein | 20 | IgG | The colour intensity of the test line is analysed using the reference score card. | Binary | The presence of a control line indicates the test is valid. | A result is positive if there is both a test line and a control line, whilst a result is negative if only the control line is present. | Using the reference score card; Positive scores ≥1 Negative scores=0 |

**Table S2: Pandemic participant laboratory-based SARS-CoV-2 antibody result.**

Breakdown of individual immunoassay results or result by one or more test.

| Test | Positive (%) | Borderline (%) | Negative (%) |
|---|---|---|---|
| **Abbott** | 310/657 (47.2%) | n/a | 347/657 (62.8%) |
| **EuroImmun** | 346/657 (52.7%) | 20/657 (3.2%) | 291/657 (44.4%) |
| **Roche** | 380/657 (57.8%) | n/a | 277/657 (42.2%) |
| **One or more test** | 385/657 (58.6%) | 3/657 (0.45%) | 269/657 (40.9%) |

**Table S3: Analytical specificity analysis on the AbC-19 LFIA** LFIAs were assessed using 34 serum samples with known other respiratory viruses, negative results for all suggests analytical specificity for SARS_CoV_2 IgG.

| SAMPLE | Number of samples | Number of AbC-19 Positive results | Number of AbC-19 Negative results |
|---|---|---|---|
| H5N1 Influenza (NIBSC 7/150) | 1 | 0 | 1 |
| RSV (NIBSC 16/284) | 1 | 0 | 1 |
| Influenza B (NIBSC 9/222) | 1 | 0 | 1 |
| Bordetella Pertussis (NIBSC 89/530) | 1 | 0 | 1 |
| Influenza A | 5 | 0 | 5 |
| Influenza B | 5 | 0 | 5 |
| Respiratory syncytial virus | 5 | 0 | 5 |
| Haemophilus Influenzae | 5 | 0 | 5 |
| Seasonal coronavirus NL63 | 5 | 0 | 5 |
| Seasonal coronavirus 229E | 5 | 0 | 5 |

**Table S4: AbC-19 LFIA results with NIBSC external reference samples**

NIBSC standard serology samples were provided with a data sheet indicating the SARS-CoV-2 antibody levels. We measured SARS-CoV-2 antibody levels in these samples and obtained similar results with the EuroImmun IgG ELISA in our laboratory.

| NIBSC # | AbC-19 LFIA result | Ulster University lab result | NIBSC provided antibody data | | | | |
|---|---|---|---|---|---|---|---|
| | | EuroImmun IgG (S1 domain) | EuroImmun IgG (S1 domain) | EuroImmun IgA | In-house IgG S1 | In-house IgG N | In-house IgG sSpike |
| **20/120** | pos (10) | pos (8.39) | pos (8.59) | pos (10.1) | 5580 | 3417 | 2693 |
| **20/122** | pos (7) | pos (3.49) | pos (3.47) | pos (1.1) | 3202 | 2425 | 1488 |
| **20/124** | pos (1) | pos (1.56) | pos (1.62) | pos (1.84) | 1636 | 3296 | 118 |
| **20/126** | pos (1) | neg (0.60) | neg (0.64) | pos (1.63) | 1181 | 995 | 8 |
| **20/128** | neg (0) | neg (0.23) | neg (0.21) | neg (0.02) | <50 | <50 | <50 |
| **20/130** | pos (8) | pos (6.96) | pos (7.77) | pos (9.74) | 5388 | 17197 | 2707 |

**Supplementary Methods**

*Laboratory-based immunoassays*

Researchers were blinded to other test results when processing these assays.

EuroImmun Anti-SARS-CoV-2 ELISA-IgG (EuroImmun, EI 2606-9601 G) was carried out according to manufacturer's instructions. Optical density (OD) at 450nm and reference OD at 620nm was read on BMG Labtech Fluostar Omega spectrophotometer (BMG Labtech). Ratios were calculated by dividing absorbance of the clinical sample by the absorbance of EuroImmun calibrator, with a score of < 0.8

determined negative, ≥ 0.8 to <1.1 borderline and ≥ 1.1 positive. For a portion of samples provided by NIBTS, EuroImmun IgG assay data was provided to researchers by NIBTS.

Roche Elecsys immunoassay (Roche Diagnostics, kit 09203079190) was carried out according to manufacturer’s instructions on the Roche cobas e601 (C6000 line) or e801 (C8000 line) analysers. The analyser automatically calculates the cut-off based on the measurement of ACOV2 Cal1 (negative) and ACOV2 Cal2 (positive). The result of a sample is given either as reactive or non-reactive as well as in the form of a cut-off index (COI; signal sample/cut-off). A score of <1.0 is determined negative, while a score ≥ 1.0 is positive.

Abbott Architect SARS-CoV-2 immunoassay was carried out according to manufacturer’s instructions on the Abbott Architect i2000SR analyser (Abbott, kit 18115FN00, calibrator kit 17412FN00, Control kit 17531FN00). The external control is entered into a Quality Monitor programme and must be within 3 standard deviations of the mean (cumulative; External control NIBSC QCRSARSCoV-2QC1 Lot 20/B764-01). Results are reported by dividing the sample result by the calibrator result. The result unit for the SARS-CoV-2 IgG assay is Index (Sample/Calibrator). A ratio of < 1.4 is determined negative and ≥ 1.4 is determined positive.

*Analytical specificity and sensitivity assessment*

Four virology samples (H5N1 influenza serology 7/150, RSV serology 16/284, Influenza B 9/222 and Bordetella Pertusis 89/530) were obtained from NIBSC (National Institute for Biological Standards, Herts, UK). An additional 30 serology samples from known virus infections were a kind gift from SugenTech, Soeul, Korea. 15 of these virology samples were obtained from Trina (Trina Bioreactives AG, Switzerland) from 5 different individuals per virus (Influenza A IgG, Influenza B IgG and RSV IgG). A further 15 of these virology samples were obtained from AbBaltris, Kent from 5 different individuals per virus (Haemophilus Influenza IgG, Seasonal Coronavirus NL63 and 229E Seasonal Coronavirus). All these serology samples alongside a panel of 6 external standard research reagents (Table S4; NIBSC; Cat: 20/118 and 20/130) were assessed on the AbC-19 LFIA to confirm analytical specificity and sensitivity.
